# Role of Growth factors (HB-EGF, VEGF-A) and Immunotolerance Mediators (TNF-αIFN-γ,PD-L1 and IL-10) in Acute and Chronic Otitis Media

**DOI:** 10.1101/2020.09.16.20195610

**Authors:** M. Selvakumari, D. AnandKarthikeyan, R. Ramakrishnan, Melvin George

**Author notes:** <u>**Address for Correspondence**:</u> Dr.Melvin George, MD, DM, Assistant Professor, Department of Clinical Pharmacology, SRM Medical College Hospital and Research Centre, SRM, University, Kattankulathur, Kancheepurram- 603203, Tamil Nadu, India.

## Abstract

**Objectives:** There is an increasing evidence of immune mediated mechanism in the etiopathogenesis of Otitis Media – ‘middle ear inflammation’. The aim of the present study was to determine the expression of important circulatoryregulators of‘immunotolerance’as biomarkers.

**Materials and Methods:** In this cross sectional study, a total of 44 OM patients and 37 controls were included. Blood plasma level of HB-EGF, IL-10, TNF-α, IFN-γ, PD-L1 and VEGF-A were quantified using Human Magnetic Luminex assay.

**Results:** The study showed statistical significant differences in the levels of VEGF-A between OM patients with and without tympanic membrane perforations (p<0.05). Moreover, we found comparatively higher level of PD-L1 in OM patients than controls. However, the level of growth factor HB-EGF, is significantly higher in controls than the cases (p<0.05). There was also a correlation between the levels of HB-EGF and VEGF-A and the severity of the disease condition.

**Conclusions:** Role of inflammatory mediators and cytokines like PD-L1, IFNγ, TNFα and IL-10in OM patients as a biomarkers are very minimal. VEFG-A have a significant role in the pathogenesis of OM.Further studies are required for better understanding of the role of these immunosuppressive mediators in the eitopathogenesis of OM.

## Introduction

“Otitis media (OM) is defined as an inflammation and infective condition of the middle ear/middleear mucosa”. If the inflammation is associated with discharge and perforation in the tympanic membrane, it results in suppurative OM. It can be acute (ASOM) or chronic (CSOM). CSOM can be further classified as squamous (safe) and mucosal type (unsafe). It is one of the most common infectious diseases of the childhood worldwide and may lead to many long-term complications, including conductive and sensorineural hearing loss (El-Sayed, 1998).Based on prevalence surveys, WHO has estimated that 28 thousand deaths occurevery year which are attributable to complications of OM. It has also been reported that hearing impairment in 42 million people (above 3 years) in the world was mainly caused by OM (Acuin, 2004). Studies around the world have reported that the prevalence of acute suppurative otitis media (ASOM) varies from 2.3% to 20%, chronic suppurative otitis media (CSOM) 4% to 33.3% and Otitis Media with Effusion (OME) from 1.3% to 31.3% (Deshmukh, 1998; Berman 1995). The prevalence rate of ASOM in India is around 17– 20%(Lazo-Saenz et al., 2005) and CSOM is 7.8% (Rupa et al., 1999).

The pathogenesis of otitis media is multifactorial which include pathogenic microbial infection, impaired eustachian tube function, recurrent upper respiratory infection, cleft palate, and adenoid hypertrophy, genetic predisposition, immature immune status, allergy, products of gastric secretion, demographic, environmental (like day care, lack of breast feeding, exposure to smoking), etc. (Usonis et al., 2016). Central to the formation of inflammation are the inflammatory mediators, which include proteins, peptides, glycoproteins, cytokines, arachidonic acid metabolites (prostaglandins and leukotrienes), growth factors, nitric oxide and oxygen free radicals (Juhn et al., 2008). Inflammation in the middle ear during infection is promoted either by increasing the proinflammatory cytokines or suppressing the anti inflammatory cytokines. Immune cells have been shown to produce inflammatory cytokines like Tumor necrosis factor alpha (TNF-α),interferon gamma(IFNγ),interluekin (IL)IL-β, IL-1, IL-4, IL-5, IL-6, IL-10 and IL-8 at different levels in various stages of disease.

There must be *‘compromised innate or adaptive immunity’* in the middle ear mucosa or ‘immunotolerance’ in OM (Lin et al., 2014). Studies suggest that immunotolerance does occur in the middle ear to a certain degree (Moriyama, et al., 1985 Bahmad and Merchant,2007). Frequently, the middle ear is tolerant to the upper respiratory infections. In the middle ear, the major immune regulators such as TNFαand IFNγ are readily secreted in response to infections, which modulate the activity of immunity. Chronic stimuli to the middle ear mucosa may form some degree of tolerance towards infectious agents when they repeatedly appear in the middle ear and cause a low profile of inflammation or immune responses by inducing the expression of programmed death ligand-1 (PD-L1), an inducible protein which is expressed in the middle ear epithelial cells and can inhibit the innate and adaptive immunity (Dong et al., 2002;Scandiuzzi et al., 2011).

TNFα is one of the most significant inflammatory mediators in OM.TNFα induces the acute phase of the inflammatory response and release of other cytokines. IFNγ is a cytokine that is critical for innate and adaptive immunity against bacteria and viruses. In COM, IFNγ is highly up-regulated in the middle ear mucosa and plays an important role in the immune response including activation of macrophages and induction of class II major histocompatibility complex (MHC) molecule expression (Zhou, 2009). It is alsoknown as a potent regulator for the expression of PD-L1 (Lin et al., 2014). It has been reported to have an immuno-regulatory role in OM with effusion (Lasisi et al., 2009).IL-10, known as cytokine synthesis inhibitory factor, is considered an immunosuppressive regulator of acute inflammation. It is produced by monocytes, CD4+ T cells, activated CD8+ T cells, and activated B cells. It is produced relatively late when compared to other cytokines. If IL-10 is present for an extended period in the inflammation zone it can induce the amplification of chronic humoral inflammatory processes. Through this humoral inflammatory amplicationof IL-10 is thought to contribute to the switching of the infection into the chronic stage.

Moreover, the involvement of growth factors has been documented in the chronicity of OM (Palacios, 2001). Heparin-binding EGF-like growth factor (HB-EGF) is one such factor which issynthesized as a transmembrane protein (proHB-EGF) and can be cleaved at the plasma membrane by metaloproteinases to yield soluble HB-EGF (sHB-EGF). HB-EGF, a member of the EGF family, is involved in epithelialization in skin wound healing rather than proliferation (Shirakata et al.,2005). Recently, it was demonstrated that HB-EGF may also contribute to angiogenesis by remodelling of vascular endothelial cells (Mehta et al., 2007).Chronic inflammation and exposure to cytokines causes vascular leak, resulting in an effusion in the middle ear space. Increased vascular destabilization and neovascularization may also be a contributing factor to disease pathology in otitis media. During chronic inflammation, angiogenesis and vascular permeability were induced by cytokines such as Vascular endothelial growth factor (VEGF). The aim of the present study was to determine if the expression of important circulatoryregulators ofimmuno tolerance were altered in patients with otitis media as compared to healthy controls.

## Materials and Methods

### Study Subjects

This was a cross-sectional study performed in the Department of Clinical Pharmacology of SRM Medical College Hospital and Research Centre, India. All the study participants were recruited from Department of Otorhinolaryngology (ENT), SRM Medical College Hospital and Research Centre, Kattankulathur and BEENT Hospital, Chengalpattu, India, and the study was conducted from February to June, 2018.

A total of 81 subjects were included based on the inclusion/exclusion criteria.Patients diagnosed with more than one episode of middle ear inflammation and infection were included in the study.The diagnosis of otorhinolaryngological problems was confirmed by ENT specialists using clinical history, clinical examinations including otoscopy, tympanometry, myringotomy and pure tone audiometry. Audiometric threshold of hearing loss was evaluated using pure tone audiometry and the average for the frequencies 0.5, 1, 2, 4 and 8 kHz was recorded. Patient history focused on age, age at onset, the number of past AOM or COMepisodes, their duration, list of antibiotics taken, family history of hearing loss, family history of otitis media and other important risk factors. Thesebase line characteristics are summarized in Table1. Patients with autoimmune diseases or taking medications for autoimmune disease for more than three months were excluded from the study. A total of 37 apparently healthy volunteers who didn’t have any otological complaints for the past six months were included in the study as controls.

**TABLE I.** Distribution of basic and clinical characteristics including demographic, risk factors, symptoms among the male and females of the study subjects

|  | **MALE**<br>**n=27** | **FEMALE**<br>**n=17** | **TOTAL**<br>**n (%)** | **P VALUE** |
| --- | --- | --- | --- | --- |
| **Age at onset** |  |  |  |  |
| 1 to 20 years | 10 | 2 | 12 (27.27) |  |
| 21 to 40 years | 12 | 10 | 22 (50.0) |  |
| >41 years | 5 | 5 | 10 (22.72) |  |
| Consanguineous Marriage | 6 | 2 | 8 (18.18) | 0.381 |
| Family history of age related Hearing loss | 4 | 5 | 9 (20.45) | 0.242 |
| **Nasopharyngeal disorder** | 8 | 11 | 19 (43.18) | 0.022* |
| Sinusitis | 4 | 6 | 10 (22.72) |  |
| Allergic Rhinitis | 4 | 3 | 7 (15.90) |  |
| Tonsillopharyngitis | - | 1 | 1 (2.27) |  |
| Asthma | - | 1 | 1 (2.27) |  |
| **Metabolic Disorder** | 2 | 5 | 7 (15.90) | 0.052 |
| Diabetes | 1 | 2 | 3 (6.81) |  |
| Hyperthyroid | - | 2 | 2 (4.54) |  |
| Hypertension | 1 | 1 | 2 (4.54) |  |
| **Symptoms** | 20 | 14 | 34 (77.27) | 0.523 |
| Facial Pain and Tinnitus | 11 | 9 | 20 (45.45) |  |
| Vertigo | 5 | 3 | 8 (18.18) |  |
| Snoring | 4 | 1 | 5 (11.36) |  |
| Spondylitis | - | 1 | 1 (2.27) |  |
| Family History of Recurrent Ear Infection | 3 | 5 | 8 (18.18) | 0.125 |
| Passive/Active/Household smoking Exposure | 10 | 6 | 16 (36.36) | 0.907 |
| Episode of Upper Respiratory Tract Infection | 11 | 3 | 14(31.81) | 0.109 |
| Recurrent Ear infection in 6 months | 10 | 8 | 18 (40.90) | 0.510 |
| **Otoscopic Findings** |  |  |  |  |
| **Laterality** |  |  |  |  |
| Unilateral | 14 | 10 | 24 (54.54) |  |
| Bilateral | 13 | 7 | 20 (45.45) | 0.651 |
| Tympanic Membrane Perforation | 12 | 9 | 21 (47.72) | 0.583 |
| **Degree of Hearing Loss** |  |  |  |  |
| Normal hearing | 13 | 12 | 25(56.81) | 0.143 |
| Hearing loss (mild to severe ) | 14 | 5 | 19 (43.18) |  |
| **Types of OM** |  |  |  |  |
| AOM | 4 | 3 | 7(15.90) | 0.803 |
| CSOM | 23 | 14 | 34(77.27) |  |

n; number denotes frequency

### Ethical approval

This study was approved by the SRM MedicalCollege Institutional Ethics Committee (IEC), India(1246/IEC/2017) and all study procedureswere performed in accordance with the provisionsof the Declaration of Helsinki. The participants were informed of the study protocol and purpose of the study and their written consent was obtained.

### Experimental analysis

Two ml of blood sample was collected in heparin citrate coated vacutainerfrom study participants. After adequate centrifugation at 1500 rpm for 10 min, the plasma samples were extracted and were stored in −80°C deep freezer. The samples were subjected to Bio-Plex^®^200 Systems(Bio Rad, California, USA). By usingHuman Magnetic Luminex Assay kit (R&D Systems, Minneapolis, USA), levels of six biomarkers (TNF-à, IFNγ, IL-10, PD-L1, HB-EGF and VEGF-A) were measured in the same sample at one-time point. The assay was run according to the manufacturer’s instructions.

### Statistical analysis

The normality of the data was checked using Q-Qplots.Continuous variables were summarized as mean ±standard deviation. Categorical data were represented as n(%). Comparison between different groups were analysed using independent student-t-test. The association between continuous variable was studied using the Pearson’s rank correlation coefficient.The optimal cut-off level of the biomarkers (HB-EGF and VEGF-A) topredict the severity of disease among the cases was evaluated using the areaunder the receiver operating characteristic (ROC) curve.All statisticalanalyses were performed with SPSS softwareV16.0 (SPSS Inc., Chicago, IL, USA). All *p-*values were two-sided with a value of <0.05 was consideredstatistically significant.

## Results and Analysis

This cross sectional study included a total of 44 cases and 37 controls. Among the cases, 84.4%(n=38) were CSOM (Safe:75.5%; unsafe:8.8%) the remaining 11.1% (n=5) were AOM and 4.4. %(n=2) were ASOM. Among the 44 subjects, 27 (61.36%) were males and 17(38.63%) were females with the mean age of 28.71± 9.95. The gender distribution among the normal healthy controls(n=37) were: males:15 (40.54%); females 22 (59.45%); mean age 25.21±4.83. Wefound that there was a significant difference in the incidence of nasopharyngeal disorder between male and female cases(p<0.05) (Table-1).Tinnitus (70.45%) andvertigo (22.72%)werethe commonly observed symptoms in both males and females. Exposure to smoking, occurrence of repeated upper respiratory tract infection, recurrent ear infection, laterality and degree of hearing loss were more common in males than in females.

The concentration of various inflammatory mediators in the plasma were recorded in terms of pg/ml. Out of the six inflammatory mediators, we found that there was a significant differencein the levels of HB-EGF, (15.45±9.73*vs*.24.54 ±26.32pg/ml, p<0.05)between the cases and controls (Table-2).However, there was a slightly higher levels of PD-L1 (3.18±11.30) wasobserved in cases than the controls. We did not find even minimal level of IL-10 expression in both the cases and control groups. The blood plasma levels of TNF-α and IFN-γwere very low in both the groups.We also attempted to analyse the difference in expression of these mediators between the OM patients with tympanic membrane perforation and OM patients without tympanic membrane perforation. We have observed that there was a significant difference between these two groups with respect to VEGF-A (*p* value < 0.05) (Table-3).

**TABLE II.** Plasma concentration of important inflammatory mediators between cases and controls

| **Inflammatory Mediators (pg/ml)** | **Patients with otitis media (n=44 )** | **Healthy controls (n= 37)** | **p-value** |
| --- | --- | --- | --- |
| HB-EGF | 15.45 ± 9.73 | 24.54 ± 26.32 | **0.037\*** |
| IL-10 | 0.0 | 0.0 | 0.0 |
| TNFα | 1.16 ±2.01 | 1.09± 2.71 | 0.90 |
| IFNγ | 0.35±2.34 | 3.45 ±19.73 | 0.30 |
| PD-L1 | 3.18±11.29 | 1.16±4.95 | 0.32 |
| VEGF-A | 23.91 ±13.5 | 24.28 ± 20.25 | 0.92 |

HB-EGF; Heparin Binding Epidermal Growth Factor; IL-10: Interleukin 10; TNF:tumor necrosis factor alpha;IFNγ ;interferon gamma; PD-L1: programmed death ligand-1; VEGF:Vascular endothelial growth factor.
\* p value > 0.05

**TABLE III.**
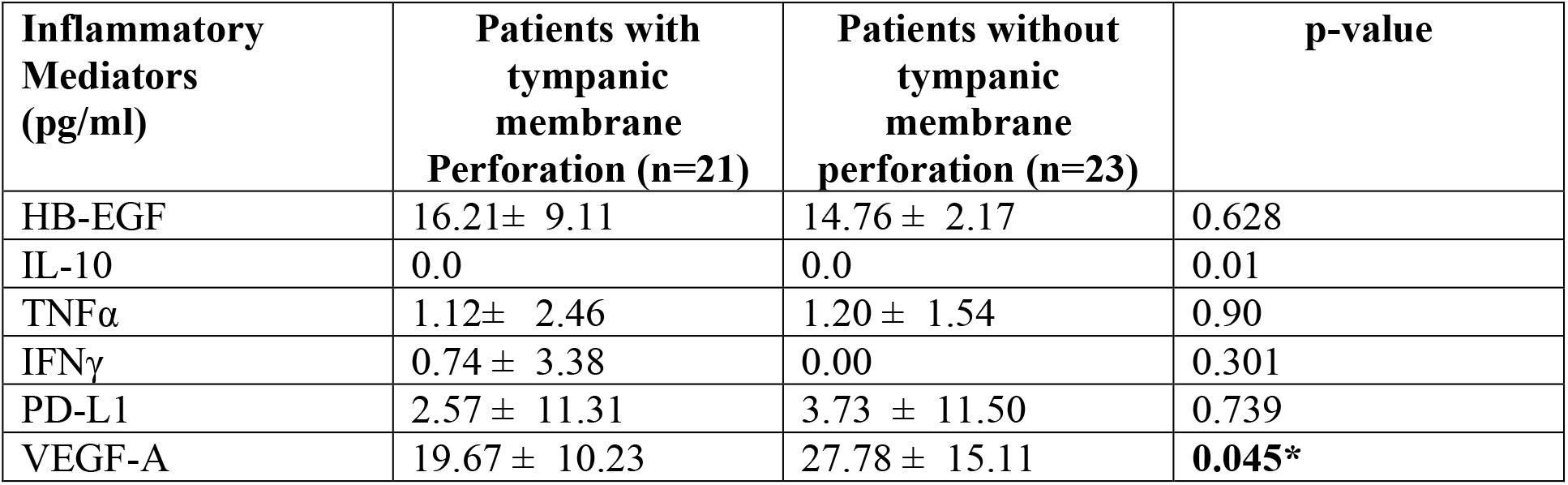
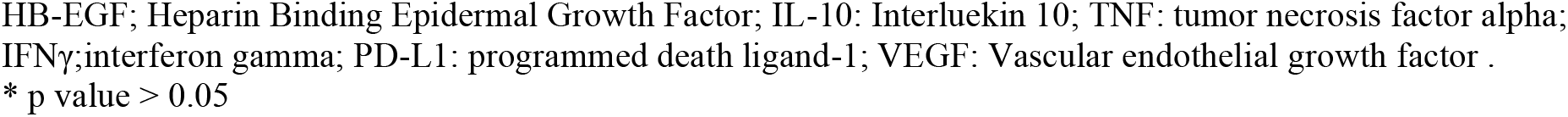
Plasma concentration of important inflammatory mediators between OM patients with tympanic membrane perforation and those without tympanic membrane perforation

| **Inflammatory Mediators (pg/ml)** | **Patients with tympanic membrane Perforation (n=21)** | **Patients without tympanic membrane perforation (n=23)** | **p-value** |
| --- | --- | --- | --- |
| HB-EGF | 16.21± 9.11 | 14.76 ± 2.17 | 0.628 |
| IL-10 | 0.0 | 0.0 | 0.01 |
| TNFα | 1.12± 2.46 | 1.20 ± 1.54 | 0.90 |
| IFNγ | 0.74 ± 3.38 | 0.00 | 0.301 |
| PD-L1 | 2.57 ± 11.31 | 3.73 ± 11.50 | 0.739 |
| VEGF-A | 19.67 ± 10.23 | 27.78 ± 15.11 | **0.045\*** |

HB-EGF; Heparin Binding Epidermal Growth Factor; IL-10; Interleukin 10; TNF: tumor necrosis factor alpha; IFNγ ;interferon gamma; PD-L1: programmed death ligand-1; VEGF: Vascular endothelial growth factor . \* p value > 0.05

The combined ROC curve was plotted to predict the disease severity. However, the study results failed to determine the cut off value for predicting the disease severity (Fig-3).

## Discussion

### Demographic Factors

The higher frequency of nasopharyngeal disorder and ear discharge seen in females could be due to frequent head bath which might have led to increased occurrence of sinusitis (Laudien, 2015). The National Health Interview Survey (2010) had reported that females accounted for 63% of sinusitis. Similarly, female preponderance for sinusitis was reported from a large population study of Olmstead County (Shashy et al., 2004).The risk factors like smoke and dust exposure were reported high in males. Similarly, unilateral or bilateral hearing loss were observed more in males whichcould be due to their exposure to noise, dust or other environmental factors. The high occurrence of CSOM among males showed their ignorance about the severity of the disease or lack of time to visit the hospital for immediate treatment.Many epidemiological studies havealso reported male preponderance in OM indicatinga greater amount of occupational and environmentalexposures(Bluestone et al., 1992; Kumar et al., 2012; kumari et al., 2016).

Interestingly, we found that eight subjects had family history of recurrent middle ear infection/inflammation for more than five years either in maternal or paternal side. This gives a clue on genetic association/predisposition to OM. There are many reports showing association of genes/genetic factors with OM (Lee et al., 2013; Esposito et al., 2014; MacArthur et al., 2014;Einarsdottir et al., 2016)

### Biomarker analysis

Previous reports showed the role of inflammatory cytokines and inflammatory mediators in the middle ear infection (Juhn et al., 2008). Some of these cytokines and inflammatory mediators are involved in the regulation of immune and inflammatory responses. Hence, the present study focuses on inflammatory mediators, HB-EGF, IFNγ, TNFα, IL-10, PD-L1 and VEGF-A and its role in the etiopathogenesis of OM

In the present study, we evaluated the blood plasma levels of HB-EGF and VEGF-A to look for its role in acute and chronic condition of OM. We found that there were significantly lower levelsof HB-EGF in cases when compared to controls. HB-EGF is one of the important growth factor that plays an important role in OM hyperplasia and its chronicity (Suzukawa et al., 2014). Studies have also shown that HB–EGF-sheds EGF ligands and regenerates chronic perforations in mouse models (Santa Maria et al., 2015),thus showing its role in healing process. This was further supported by another study on acute wound healing of the tympanic membrane (Santa Maria et al., 2011). Lower expression rate of HB-EGF in OM patients suggest that healing process in the middle ear inflammation is very minimaland thus leads to severity of the pathogenesis of OM.

VEGF is another growth factor important for vascular permeability and angiogenesis. Though in the present study we did not find any significant differences in levels of VEGF between cases and controls, we could see that there was a significant difference between the OM patients with tympanic membrane perforation and patients without tympanic membrane perforation. Recurrent cytokine exposure during inflammation, leads to vascular leak causing effusion or fluid accumulation in the middle ear mucosa. Increased VEFG expression has been observed during neovascularisation and eustachian tube dysfunction (Lim and Birck, 197; Ryan 1993; Huang et al.,2012). VEGF expression was identified in effusion fluid and middle ear mucosa of human patients with otitis media (Jung et al., 1999).Another study found VEGF protein expression in themiddle ear effusions of all the33 pediatric patients (Sekiyama et al., 2011).This observation demonstrates that the role of VEFG-A in neovascularisation and in middle ear effusion.

In addition to the above, this study also focuses on the inflammatory mediators (TNFα, IFNγ, IL-10 and PD-L1) in the immunotolerance or peripheral tolerance occurringin the middle ear. We could find low levels of these mediators and there were no significantdifferences between the groups, implies that the role of these mediators in OM condition would be minimal or organ specific.

One of the proposed hypothesis for the chronic or recurrent OM is immunotolerance or compromised innate or adaptive immunity (Alford et al., 1976). Duringthe acute condition, the middle ear epithelial cells starts producing first level cytokines in response to pathogenic infection via the NF-κBsignalingpathway (Li, et al., 2002; Rhodus et al., 2005). At the chronic stage, secretion of IFNγ and TNFα regulates the expression of Programmed death-ligand 1 (PD-L1),a 40kDa type 1 transmembrane protein, speculated to play a major role in immune suppression by reducing the proliferation of antigen-specific T-cells and reducing apoptosis in regulatory T cells (Schoenborn et al., 2007; Lin 2014).This action leads to the formation of the immune tolerance towards the pathogenic invasion. In this study though we couldn’t find any significant differences in the value of PD-L1, we found higher levels of PD-L1 in cases than the controls. This observation may suggest that some amount of immunotolerance activity was performed in the middle ear mucosa.

IL-10, known as cytokine synthesis inhibitory factor, is considered as an immunosuppressive regulator of acute inflammation. It is produced relatively late when compared to other cytokines and suppressing the inflammatory reactions by controlling the production of proinflammatory cytokines.Through the humoral inflammatory amplification, IL-10 is thought to contribute to the switching of the infection into the chronic stage. Hence, absence of IL-10, low levels of PD-L1 IFNγ, TNFα in this present study may suggests that these mediators of immuno tolerance may not be considered as a predictor marker for late stages of OM progression. The chances of these marker expression might be high during early stages of chronic condition or when genetic predisposition exists (Lin et al., 2014). Another possible reason could be samples used in this study may have low levels of circulatory inflammatory mediators when compared to middle ear effusion fluids.

### Study Limitations

The main limitations of our study is with respect to smaller sample size.The samples were collected at different points of time and the baseline study medications of the patients were not recorded, which could have an effect on biomarkers concentration. Additionally, no follow up of the study subjects was done.

## Conclusion

In conclusion, our study showed that the role of mediators and cytokines likePD-L1, IFNγ, TNFα and IL-10in OM patients as a biomarkers are very minimal. However,role of these mediators at the site of infection and in directing the progression of the diseases cannot be ruled out. Only limited studies are available on effect of growth factors on the hyperplastic response of the middle ear mucosa. The different levels of growth factors (HB-EGF, VEGF-A) seen in the present study signifies its potential role inthe pathophysiology of Otitis Media. An enhanced understanding of the role of these growth factors could help us to identify better strategies in targeting therapies towards OM.Thus, future prospective cohort studies are needed to confirm our findings.

## Data Availability

The data referred to in the manuscript are original includes tables and figures

## Acknowledgements

We deeply acknowledge the co-operation extended by all the patients during this study. We wish to acknowledge the Department of Clinical Pharamacology, SRM Medical College Hospital and Research Centre, for providing laboratory support. We thank Ms.LuxitaaGoenka for statistical analysis. We thank Dr.SundarRajan, ENT, BEENT Hospital, Chengalpattu for extending his support to provide the samples. We acknowledge the Bioplex facility provided by SRM-DBT Platform for Advanced Life Science Technologies, Kattankulathur. M.Swas supported by a Research Associate Fellowship (2017-2018) from the Department of Biotechnology. This study was partly supported by DBT-RA-Contingency grant of M.S.

## Declaration of Statement

### SUMMARY

- There is *‘compromised innate or adaptive immunity’* in the middle ear mucosa or ‘immunotolerance’ in OM. The present study is the first attempt to determine if the expression of important circulatory regulators of immuno tolerance (HB-EGF, IL-10, TNF-α, IFN-γ, PD-L1 and VEGF-A) were altered in patients with otitis media with chronicity.
- The study includes a total of 44 cases and 37 controls. Among the cases, 84.4%(n=38) were CSOM (Safe:75.5%; unsafe:8.8%) the remaining 11.1% (n=5) were AOM and 4.4. %(n=2) were ASOM.
- Wefound that there was a significant difference in the incidence of nasopharyngeal disorder between male and female cases(p<0.05).Tinnitus (70.45%) andvertigo (22.72%)werethe commonly observed symptoms in both males and females.
- We found that there was a significant differencein the levels of HB-EGF, (15.45±9.73*vs*.24.54 ±26.32pg/ml, p<0.05)between the cases and controls
- There was a significant difference between the groups with and without tympanic membrane perforation in terms of VEGF-A (*p* value < 0.05)
- Thus an enhanced understanding of the role of these growth factors could help us to identify better strategies in targeting therapies towards OM. Nevertheless, future prospective cohort studies are needed to confirm our findings.

---

**FIGURE I.**
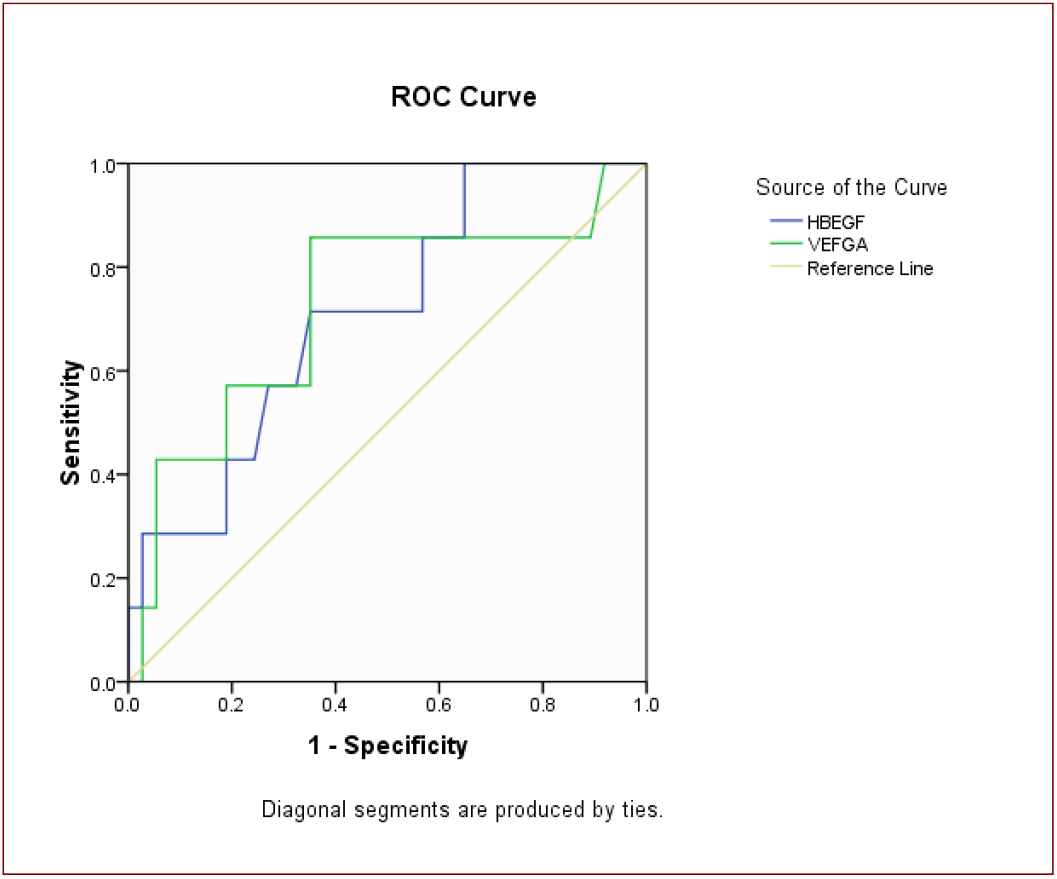
ROC curve for prediction of severity of Otitis Media. Combined receiver-opening characteristic (ROC) curves of HB-EGF and VEGF-A to predict the severity of disease. AUC = 0.710; *p* = 0.080; 95% CI (0.515–0.906). AUC = 0.724; *p* = 0.116; 95% CI (0.496–0.952). HB-EGF Heparin Binding Epidermal Growth Factor; VEGF-A;Vascular Endothelial Growth Factor-A AUC, area under the curve; CI, confidence interval

## References

Acuin J. Chronic suppurative otitis media - Burden of Illness and Management Options. Geneva: World Health Organization; 2004.

Alford BR, McFarlane JR, Neely JG. Homograft replacement of the tympanic membrane. Laryngoscope 1976;86:199–208

Bahmad F Jr, SN Merchant. Histopathology of ossicular grafts and implants in chronic otitis media. Ann OtolRhinolLaryngol 2007; 116: 181–91

Berman S. Otitis media in developing countries. Pediatrics 1995;96:126–31.

Bluestone CD, Stephenson JS, Martin LM. Ten-year review of otitis media pathogens. Pediatr Infect Dis J 1992;11: (Suppl) 7–11

Deshmukh CT. Acute otitis media in children-treatment options. J Postgrad Med 1998;44:81–4.

Dong H, Strome SE, Salomao DR, Tamura H, Hirano F, et al. Tumor-associated B7-H1 promotes T-cell apoptosis: a potential mechanism of immune evasion. Nat Med 2002;8: 793–800

Einarsdottir E, Hafrén L, Leinonen E, Bhutta M F, Kentala E, Kere J and Mattila PS. Genome-wide association analysis reveals variants on chromosome 19 that contribute to childhood risk of chronic otitis media with effusion. ScienRep; 2016; 6:33240 :1–11

El-Sayed Y. Bone conduction impairment in uncomplicated chronic suppurative otitis media. Am J Otolaryngol. 1998;19:149–53

Esposito S, Marchisio P, Orenti A, Spena S, Bianchini S, Nazzari E, Rosazza C, Zampiero A, Biganzoli E, Principi N; Genetic polymorphisms of functional candidate genes and recurrent acute otitis media with or without tympanic membrane perforation. Medicine 2015; 94(42):e1860.

Huang Q, Zhang Z, Zheng Y, et al. Hypoxia-inducible factor and vascular endothelial growth factor pathway for the study of hypoxia in a new model of otitis media with effusion. Audiol Neurootol 2012;17:349–56.

Juhn SK, Jung MK, Hoffman MD, Drew BR, Preciado DA, et al. The role of inflammatory mediators in the pathogenesis of otitis media and sequelae. ClinExpOtorhinolaryngol 2008;1: 117–38.

Jung HH, Kim MW, Lee JH, et al. Expression of vascular endothelialgrowth factor in otitis media. ActaOtolaryngol 1999;119:801–08.

Kumar N, Chilke D, Puttewar MP. Clinical profile of tubotympanic CSOM and its management with special reference to site and size of tympanic membrane perforation, eustachian tube function and three flap tympanoplasty. Indian J Otolaryngol Head NeckSurg. 2012;64:5–12.

Kumari MS, Madhavi J, Krishna N B, Meghanadh K R, Jyothy A. Prevalence and associated risk factors of otitis media and its subtypes in South Indian population. Egypt J Ear, Nose, Throat and Allied Sci 2016;17: 57–62

Lasisi AO, Olayemi O, Arinola OG, Omilabu SA, et al. Interferon-gamma in suppurative otitis media:significance of otorrhoea type and disease outcome. J LaryngolOtol 2009; 123: 1103–07.

Laudien M. Orphan diseases of the nose and paranasal sinuses: Pathogenesis– clinic therapy. GMS Curr Top Otorhinolaryngol Head Neck Surg 2015:14, Doi :10.3205/cto000119

Lazo-Saenz JG, Galvan-Aguilera AA, Martínez-Ordaz VA, Velasco-Rodríguez VM,. Nieves-Rentería A, Rincon-Castaneda C. Eustachian tube dysfunction in allergic rhinitis. Otolaryngol Head Neck Surg, 2005; 132: 626–629

Lee HY, Chung JH, Lee SK, et al. Toll-like receptors, cytokines & nitric oxide synthase in patients with otitis media with effusion. Indian J Med Res. 2013;138:523– 30.

Li J, Kartha S, Iasvovskaia S, Tan A, Bhat RK, et al. Regulation of human airway epithelial cell IL-8 expression by MAP kinases. Am J Physiol Lung Cell MolPhysiol 2002; 283: L690–99.

Lim D, Birck H. Ultrastructural pathology of the middle ear mucosa in serous otitis media. Ann OtolRhinolLaryngol 1971;80:838–53.

Lin J. Basic science concept in otitis media pathophysiology, in State of the art concepts and treatment, D. Preciado (Ed); 2014.

MacArthur CJ, Wilmot B, Wang L, Schuller S, Lighthall J, Trune D. Genetic susceptibility to chronic otitis media with effusion: candidate gene SNPs Laryngoscope. 2014; 124(5): 1229–1235.

Mehta, VB, Besner GE. HB-EGF promotes angiogenesis in endothelial cells via PI3-kinase and MAPK signaling pathways. Growth Factors 2007; 25: 253–63.

Moriyama H, Aoki K, Honda Y. Homografts of the tympanic membrane with malleus; histological study in cat. AurisNasus Larynx 1985;12: 73–80

Palacios SD, Oehl HJ, Rivkin AZ, Aletsee C, Pak K, Ryan AF. Growth factors influence growth and differentiation of the middle ear mucosa. Laryngoscope 2001;111: 874–80.

Rhodus NL, Cheng B, Myers S, Bowles W, et al. A comparison of the pro-inflammatory, NF-kappaB-dependent cytokines: TNF-alpha, IL-1-alpha, IL-6, and IL-8 in different oral fluids from oral lichen planus patients. ClinImmunol 2005;114: 278–83.

Rupa V, Jacob A, Joseph A. Chronicsuppurative otitis media: prevalence and practices among rural South Indian children. Int J PediatrOtorhinolaryngol 1999;25;48(3):217–21

Ryan AF, Baird A. Growth factors during the proliferation of the middle ear mucosa. ActaOtolaryngol 1993;113:68–74.

Santa Maria PL, Redmond SL, McInnes RL, Atlas MD, Ghassemifar R. Tympanic membrane wound healing in rats assessed by transcriptome profiling. Laryngoscope 2011;121(10), 2199–213

Santa Maria PL, Kim S, Varsak YK, Yang YP. Heparin binding–epidermal growth factor-like growth factor for the regeneration of chronic tympanic membrane perforations in mice. Tissue En g 2015; 21(9-10): 1483–94

Scandiuzzi L, K Ghosh, X Zang. T cell costimulation and coinhibition:genetics and disease. Discov Med 2011;12: 119–28.

Schoenborn JR, CB Wilson. Regulation of interferon-gamma during innate and adaptive immune responses. AdvImmunol 2007; 96: 41–101.

Sekiyama K, Ohori J, Matsune S, Kurono Y. The role of vascular endothelial growth factor in pediatric otitis media with effusion. AurisNasus Larynx 2011;38:319–24.

Shashy RG, Moore EJ, Weaver A. Prevalence of the chronic sinusitis diagnosis in Olmsted County, Minnesota. Arch Otolaryngol Head Neck Surg 2004;130:320–23.

Shirakata Y, Kimura R, Nanba D, Iwamoto R, Tokumaru S, Morimoto C, et al. Heparin-binding EGF-like growth factor accelerates keratinocyte migration and skin wound healing. J Cell Sci. 2005;118(Pt 11):2363–70.

Suzukawa K, Tomlin J, Pak K, Chavez E, Kurabi A, Baird A, Wasserman SI, Ryan AF. A mouse model of otitis media identifies HB-EGF as a mediator of inflammation-induced mucosal proliferation. Plos One 2014; 9 (7): e102739

Usonis V, Jackowska T, Petraitiene S, Sapala A, Neculau A, Stryjewska I, et. al. Incidence of acute otitis media in children below 6 years of age seen in medical practices in five East European countries BMCPediat 2016;16:108

Zhou F. Molecular mechanisms of IFN-gamma to up-regulate MHC class I antigen processing and presentation. Int Rev Immunol 2009;28: 39–60.

